# Predicting brain amyloid-β PET phenotypes with graph convolutional networks based on functional MRI and multi-level functional connectivity

**DOI:** 10.1101/2021.08.26.21262325

**Authors:** Chaolin Li, Mianxin Liu, Jing Xia, Lang Mei, Qing Yang, Feng Shi, Han Zhang, Dinggang Shen

**Affiliations:** Experimental Center, Guangzhou University, Guangzhou, China; School of Biomedical Engineering, ShanghaiTech University, Shanghai, China; Institute of Brain-Intelligence Technology, Zhangjiang Lab, Shanghai, China; Department of Research and Development, United Imaging Intelligence Co., Ltd., Shanghai, China

**Keywords:** Functional connectivity, brain network, amyloid β, PET, graph convolutional network

## Abstract

The detection of amyloid-β (Aβ) deposition in the brain provides crucial evidence in the clinical diagnosis of Alzheimer’s disease (AD). However, the efficiency of the current PET-based brain Aβ examination suffers from both coarse, visual inspection-based bi-class stratification and high scanning cost and risks. In this work, we explored the feasibility of using non-invasive functional magnetic resonance imaging (fMRI) to predict Aβ-PET phenotypes in the AD continuum with graph learning on brain networks. First, three whole-brain Aβ-PET phenotypes were identified through clustering and their association with clinical phenotypes were investigated. Second, both conventional and high-order functional connectivity (FC) networks were constructed using resting-state fMRI and the network topological architectures were learned with graph convolutional networks (GCNs) to predict such Aβ-PET phenotypes. The experiment of Aβ-PET phenotype prediction on 258 samples from the AD continuum showed that our algorithm achieved a high fMRI-to-PET prediction accuracy (78.8%). The results demonstrated the existence of distinguishable brain Aβ deposition phenotypes in the AD continuum and the feasibility of using artificial intelligence and non-invasive brain imaging technique to approximate PET-based evaluations. It can be a promising technique for high-throughput screening of AD with less costs and restrictions.

## 1. Introduction

Alzheimer’s disease (AD) is a serious progressive neurodegeneration disease. It causes increasing challenges to the public health system worldwide. In 2020, there were about 50 million AD patients in the world, and this number may increase to 152 million by 2050 (WHO, 2020). It is estimated that the global expenditure caused by AD will reach $9.12 trillion in the near future (Jia et al., 2018). Mild cognitive impairment (MCI) is regarded as an intermediate stage between normal cognition to AD. In 2011, the National Institute on Aging-Alzheimer’s Association workgroups proposed to use “continuum” to describe the whole development process from normal cognition to MCI and then AD (Sperling et al., 2011), with official term “AD continuum” used in 2018 (Jack, C.R. et al., 2018).

Disproportionate amyloid-β (Aβ) deposition in the brain has been regarded as a sensitive biomarker to detect AD at its early stage such as MCI (McKhann et al., 2011). Recent studies have highlighted positron emission tomography (PET) as a key *in vivo* technology to detect brain Aβ deposition (Müller et al., 2019; Shi et al., 2020; Zammit et al., 2021). Two types of PET are clinically prevalent: fluorodeoxyglucose (FDG)-PET that measures the glucose metabolism of the brain, an indirectly measurement of abnormalities in brain activity, often treated as a supplementary diagnostic technique in AD, and Aβ-PET that directly measures spatial deposition of Aβ in the brain, clinically regarded as “gold standard” in AD diagnosis. In Aβ-PET, the Aβ-sensitive radiotracer injected beforehand will concentrate near Aβ, and PET scan assesses the uptake value of the Aβ radiotracer in the brain, often called standard uptake value (SUV).

However, two limitations have hindered wider clinical applications of the Aβ-PET in early AD diagnosis. *First*, the clinical report of Aβ-PET scans for AD diagnosis often constitutes binary results, i.e., clinicians often stratify Aβ-PET phenotypes to either negative or positive one. Such decision making is neither operative nor objective, largely limited by the visual inspection and the experience of diagnosticians. Diagnosis bias or false positives/negatives are unneglectable. *Second*, such a rough, binarized assessment may not be suitable for characterizing the AD continuum, where the different stages need to be described with carefully designed grades. There could be distinguishable spatial patterns of brain Aβ-PET associated with different AD stages. So far, few studies have explored the brain Aβ-PET patterns searching for informative and distinctive imaging phenotypes (Wolk et al., 2009; Laforce & Rabinovici, 2011).

Besides that, PET scan are not as commonly equipped as MRI scanners in general hospitals, partly due to the fact that PET scanner and PET imaging are expensive, time-consuming, with many clinical and environmental restrictions, such as the restriction to be performed on patients with renal insufficiency. Taking together, PET it not suitable for a large-scale early AD screening for the much larger risk populations. Recent studies have suggested that non-invasive MRI techniques, such as resting-state functional MRI (fMRI), could be potentially useful for early AD detections (Ju et al., 2019), mostly using the derived functional connectivity (FC) as imaging markers of AD (Kam, T.E. et al., 2020; Li, Y. et al., 2020). Quantifying temporal correlations among the Blood Oxygen Level Dependent (BOLD) signals in different brain regions, previous FC studies had found that Aβ deposition in the brain might lead to FC alterations that can be detected by fMRI (Hedden et al., 2009; Elman et al., 2016). This inspired the current study to assess Aβ-PET phenotypes by predicting Aβ-PET grading with non-invasive fMRI and brain FC networks.

With methodological development of FC, dynamics-based high-order functional connectivity (dHOFC) (Chen et al., 2016) has been applied in early AD detection studies with fMRI, providing supplementary and better sensitivity to FC-based detection (Lei et al., 2021). To better learn the representations of brain networks, deep graph learning with graph convolutional network (GCN) (Kipf & Welling, 2016a) has been proposed in AD fMRI studies with better detection accuracy than traditional machine learning. Hence, we proposed using fMRI and the derived FC and dHOFC networks to approximate Aβ-PET-based diagnosis with newly the proposed GCN, to facilitate future high-throughput screening of AD with less costs and restrictions. Unlike most of the previous studies, we did not use neurophysiological tests- or questionnaires-based diagnostic labels (such as MCI), but instead using Aβ-PET *per se* to generate “ground truth Aβ phenotypes” for our computer-aided diagnostic modeling. We argue that the diagnosis of brain Aβ phenotypes may be more clinically desired than cognitively-defined MCIs in early AD diagnosis. The results showed that it is feasible to use advanced brain network modeling and machine learning on resting-state fMRI as a non-invasive and low-cost approximation to Aβ-PET phenotyping for future AD screening. The novelty of this study is three-fold: 1) we identified three typical brain Aβ phenotypes with meaningful associations with other clinical data (gender, age, cognitive ability, and genotype), which could be related to different progression stages of the AD continuum; 2) we proved that successful prediction to Aβ-PET patterns instead of cognitive performance-based labels can be achieved using fMRI; 3) we demonstrated the effectiveness of GCN framework in learning multi-level topological information from the brain functional connectome toward AD phenotyping.

## 2. Materials and Methods

### 2.1 PET and fMRI data collection

We used the Open Access Series of Imaging Studies 3 (OASIS-3) dataset (LaMontagne et al., 2019) and selected 258 samples from a single imaging center with complete structural T1-weighted MRI, resting-state fMRI, and Aβ-PET scans. The resting-state fMRI had 164 time points with a spatial resolution of 4 × 4 × 4 mm^3^ and a temporal resolution of 2.2 s. The PET images included 26 volumes with a spatial resolution of 2.3 × 2.3 × 2 mm^3^. The radiotracer injected was the [^11^C]-Pittsburgh compound B (PiB) for Aβ tracing. For more details of the used data, please see (LaMontagne et al., 2019). According to the conventional questionnaire-based diagnosis criteria, there were 227 samples with normal cognitive function (NCs), 14 with MCI due to various reasons, and 17 with AD. As stated above, we did not use these labels, but according to the clinical practice, using a data-driven, clustering method to identify different Aβ-PET phenotypes as the labels to be predicted with fMRI.

### 2.2 Multi-level, fMRI-based graph learning for brain Aβ-PET phenotype prediction

We used the following framework to identify the Aβ-PET phenotypes and approximate phenotypic labels with deep graph learning on the multi-level, fMRI-based brain functional connectome (Fig. 1). First, a clustering analysis was performed on the spatial patterns of the Aβ-PET images to reveal different brain Aβ phenotypes. Second, multi-level, fMRI-based brain functional connectome was constructed using Pearson’s correlation (PC, i.e., the conventional FC) and the dHOFC (Chen et al., 2016), respectively. Third, a graph convolutional learning model (GCN) was trained to jointly use PC- and dHOFC-derived brain networks for Aβ-PET phenotype prediction. Of note, all the above processing were based on the training set (part of the 258 samples) without seeing the testing set (the rest of the 258 samples), and the trained model was tested on the testing set to avoid double-dipping.

**Fig. 1.**
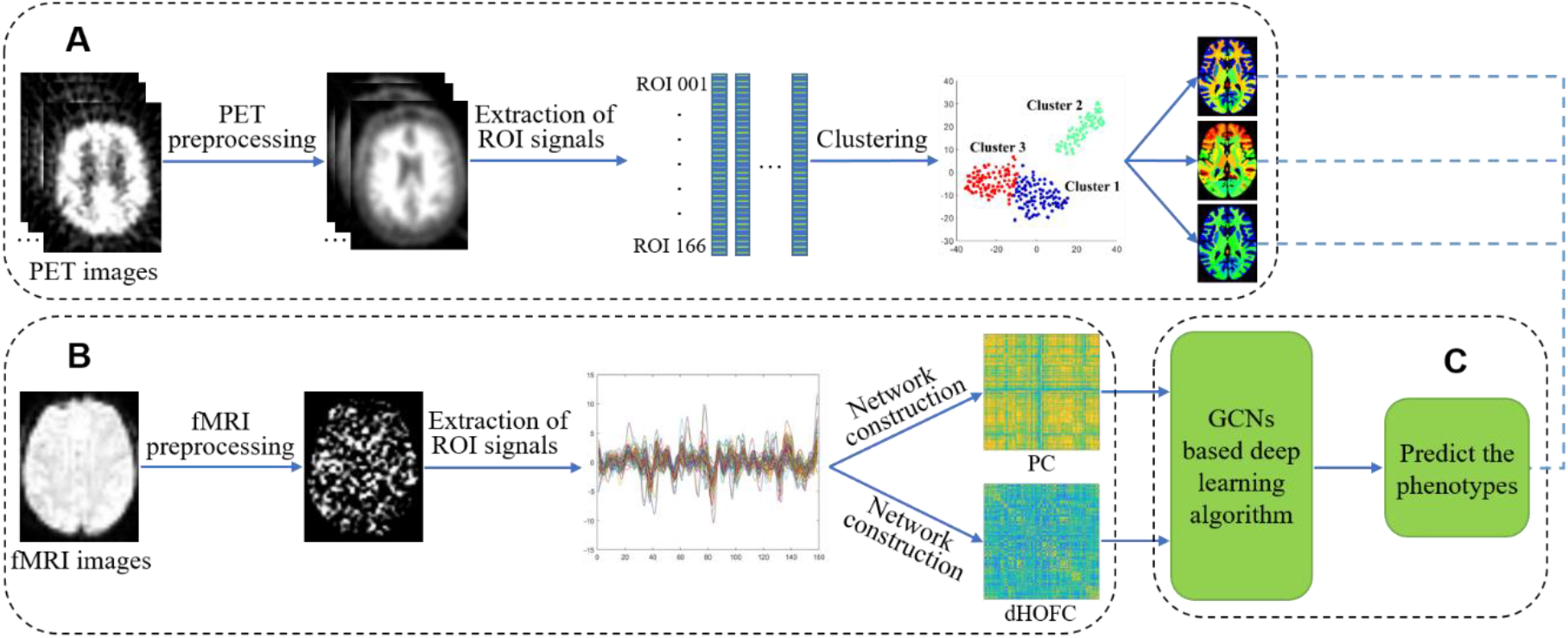
Pipeline of multi-level, fMRI-based graph learning for brain Aβ-PET phenotype prediction. It consists of three parts: A) Identifying different Aβ-PET phenotypes through a clustering analysis on the spatial pattern of the Aβ-PET images of all training samples; B) Multi-level functional connectivity (FC) networks construction with Pearson’s correlation (PC, conventional method, low-order) and dynamics-based high-order functional connectivity (dHOFC), respectively; and C) Predicting Aβ-PET phenotypes with graph convolutional networks (GCNs), where the prediction model was trained on the training set and tested on the testing set.

#### 2.2.1 Identify Aβ-PET phenotypes

PET images were preprocessed by using PET Unified Pipeline (PUP, https://github.com/ysu001/PUP) (Su et al., 2015). Briefly, the regional spread function based partial volume correction was performed to correct the SUV ratio (SUVR), where the SUVR was derived by dividing the target SUV of each voxel by the averaged cerebellar SUV. Then, the entire PET image of each sample was parcellated using the Desikan-Killiany (DK) atlas in the FreeSurfer software (Desikan et al., 2006), with each regional averaged SUVR extracted. We used 166 regions of interest (ROIs) in the DK atlas, after excluding unrelated regions, such as corpus callosum, ventricle, cerebrospinal fluid (CSF), and other unknown regions. The OASIS dataset provided the Aβ-PET SUVR data extracted from each brain region. To reduce the feature dimension from 166 to a lower one for increasing the clustering robustness, the low dimension features of the SUVR values was extracted by using the *t*-distributed stochastic neighbor embedding (*t*-SNE) (Laurens & Hinton, 2008), and then a *k*-means (Euclidean distance-based) clustering was carried out on the low-dimension *t*-SNE features to classify the dimension-reduced PET SUVR data into different phenotypes (i.e., clusters) (Mwangi et al., 2014). A total of 200 iterations were conducted to accommodate the stochastic nature of the clustering algorithm. According to the elbow method and the silhouette coefficient, the optimal cluster number was set as three for the *k*-means clustering. The three clusters were visualized on a 2D plane (see Fig. 1A) with the *t*-SNE nonlinear distance-keeping dimensionality reduction algorithm. By further checking other clinical information (i.e., age, cognitive ability, and genotype), we found that such an optimized cluster number yielded biologically meaningful characterization of the AD progression (see Result). The representative Aβ-PET phenotypes (i.e., the cluster center) derived from clustering were visualized by using the Mricron toolbox (Version 1.0.20190902, https://www.nitrc.org/projects/mricron).

#### 2.2.2 Construct multi-level FC networks based on fMRI

The resting-state fMRI data were preprocessed by Brainnetome fMRI toolkit (BRANT) 3.36 (Xu et al., 2018) (http://brant.brainnetome.org/en/latest), including removal of the first five time points, slice timing correction, head motion correction with less than 3 mm or 3°, fMRI co-registration to the associated T1-weighted structural MR images, normalization respective tissue probability images (derived from segmentation of the T1 images) to the tissue probability maps (TPM) in the standard space and applying the warp information to the fMRI data, reslicing the normalized fMRI data to 3 × 3 × 3 mm^3^, spatial smoothing with a 6 mm full-width at half-maxima (FWHM) of Gaussian kernel, temporal detrending and band-pass filtering (0.01–0.08 Hz), and covariates regression for artifact reduction. Then, we used a functional brain parcellation atlas consisting of 268 ROIs from (Finn et al., 2015) to extract regional averaged fMRI signals for FC networks construction.

Two types of FC networks were constructed using the BrainNetClass toolbox (Zhou, Z. et al., 2020) (https://github.com/zzstefan/BrainNetClass), each representing a different functional organization level in the resting brain. Here, we used two FC network construction methods that have been extensively adopted in the previous papers with one supplementing the other for better brain disease detection (Chen et al., 2016; Zhao et al., 2020). The PC-based (low-order) FC network characterizes the temporal synchronization of the BOLD fMRI signals for every pair of brain regions, while the dHOFC-based (high-order) FC network defines temporally coordinated FC dynamics among different brain region (connection) pairs (Chen et al., 2016). Therefore, the PC-based FC defined static FC between every pair of regions, while the dHOFC utilized dynamic FC and its higher-level temporal coordination among different region pairs; the two networks built reflected different levels of resting-state brain functional organization, which was hypothesized to better represent the Aβ-PET patterns. The dHOFC network construction was described elsewhere (Zhou, Z. et al., 2020) as briefly summarized in the following three steps. First, dynamic FC was calculated between each pair of the ROI signals with a sliding window (window length = 66 s, step size = 2.2 s). Second, all the dynamic FC time series was clustered into 600 clusters by using agglomerative clustering, with each mean time series of each cluster representing a dynamic FC pattern. Third, the temporal synchronization was measured for each pair of the clusters (Zhou, Z. et al., 2020).

According to the used functional atlas, we obtained a 268 × 268 PC-based FC matrix and a 600 × 600 dHOFC matrix for each sample, which were used for Aβ-PET phenotype prediction.

#### 2.2.3 Predict Aβ-PET phenotypes with graph learning on multi-level FC graphs

We proposed to use GCN, one of the most effective graph learning algorithm, to learn Aβ-PET phenotypic representations with different branches separately learning the PC- and dHOFC-based multilevel FC networks for a joint Aβ-PET phenotype prediction (Fig. 2). GCN can automatically extract topological and node features layer by layer and encode them into low dimensional representation (Kipf & Welling, 2016a). The effectiveness of the GCN in the studies of neurological disease diagnosis has been widely demonstrated in AD, MCI, and major depressive disorder diagnosis (Wee et al., 2019; Jun et al., 2020; Liu et al., 2020).

**Fig. 2.**
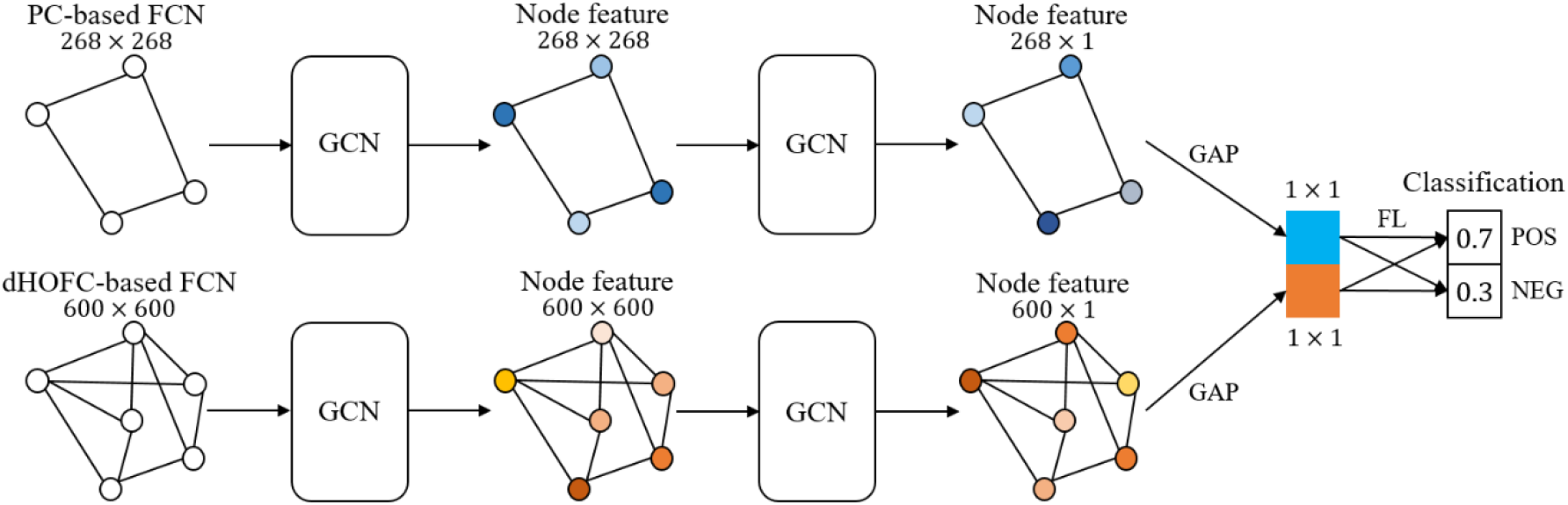
The two-branch GCN network architecture for jointly PC- (the upper branch) and dHOFC- (the lower branch) based brain functional connectome learning for a two-class Aβ-PET phenotype classification. PC: Pearson’s Correlation, dHOFC: dynamics-based High-Order Functional Connectivity, FCN: Functional Connectivity Network, GCN: Graph Convolutional Network, GAP: Global Average Pooling, FL: Fully-connected Layer, POS: Positive, NEG: Negative.

Specifically, we used GCN to learn both nodal features and topological features from two types of the connectivity graphs. For the PC-based FC network, the nodal features of the graph were defined as the one-hot code of each of the 268 brain ROIs and the topological features were defined as the FC between each pair of the brain ROIs. For the dHOFC network, the nodal and topological features were defined likewise. Of note, the topological features we used were the weighted FC or dHOFC strength without any binarization operation (i.e., weighted graph) to avoid bias of the binarization threshold or density threshold. A graph convolution was performed on each queried node of each type of the graphs to generate new nodal representations of the queried node by aggregating the nodal information from the neighboring nodes through the topological information of the queried node. We used two graph convolution layers for extraction of more diagnostic, deeper nodal features (Kipf & Welling, 2016b). Since the FC and dHOFC graphs represents different levels of brain functional organizations that could be of diagnostic importance and could supplement each other, two branches of GCN were separately constructed and their outputs (nodal features) were further integrated by concatenating each global average pooling (GAP) output, which was then forwarded to a fully-connected layer (FL) with *dropout* and *softmax* operations to obtain the final prediction results. Since three Aβ-PET phenotypes were detected using clustering and we hypothesized that different brain network features contributed differently in each pairwise Aβ-PET phenotype differentiation task, we conducted three two-class classification using the above two-branch GCN framework trained separately, for phenotype 1 vs. 2, phenotype 1 vs. 3, and phenotype 2 vs. 3.

The weighted cross-entropy was used as a loss function to solve the problem of unbalanced classifications because, in our study, the sample sizes of the three Aβ-PET phenotypes were unbalanced. Five-fold cross-validation was conducted to evaluate model performance. In each fold, we randomly split the samples into a training set (80% of the total sample size) and a testing set (20%). To avoid double-dipping, only the training set was used to conduct the clustering-based Aβ-PET phenotype identification in Section 2.2.1, while the Aβ-PET phenotype labels of the testing set were determined by majority voting from the *k*-nearest neighbors (*k* = 20) with their distance to each of the three cluster centers, where the cluster with the most votes was assigned to the test data. For quantitative comparison with the double-branch GCN framework for jointly prediction, we also conducted single-branched GCN-based Aβ-PET phenotype bi-class classifications using PC-based and dHOFC-based networks, separately. Model performance metrics, including accuracy, sensitivity, and specificity across the five folds were computed and reported.

## 3. Results

### 3.1 Aβ-PET phenotypes

According to the clustering results and the optimized cluster number, three clusters were identified and their centroids were respectively visualized as representative Aβ-PET phenotypes in Fig. 3. The 2D distance-keeping visualization of the three clusters (in different colors) was shown in Fig. 4, where the three clusters were clearly separated from each other, with the Cluster 2 singled out from two much closer clusters 1 and 3. Of note, this is for visualization purpose; therefore, all data was used. In the presented results, there were 97 samples in the Cluster 1, 74 samples in the Cluster 2, and 87 samples in the Cluster 3.

**Fig. 3.**
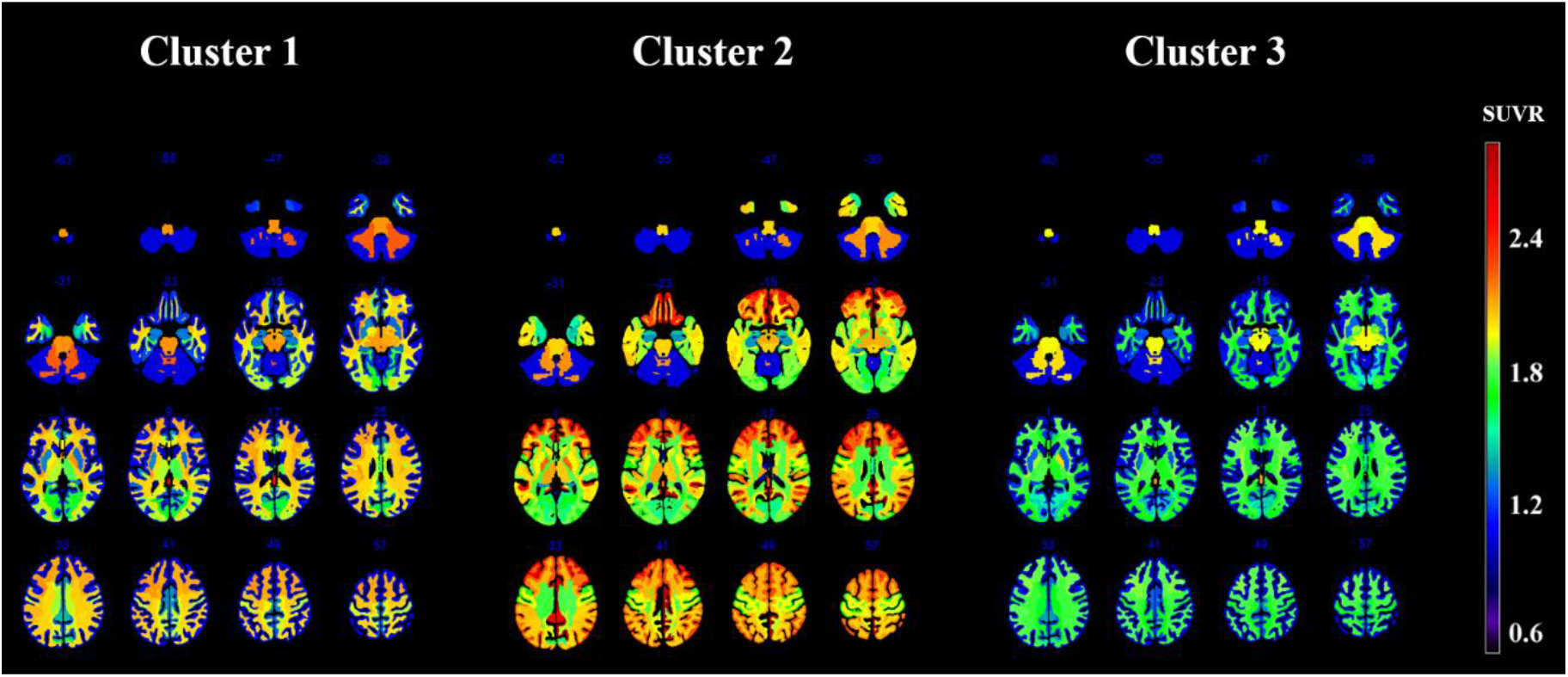
Visualizations of the representative Aβ-PET phenotypes derived from clustering.

**Fig. 4.**
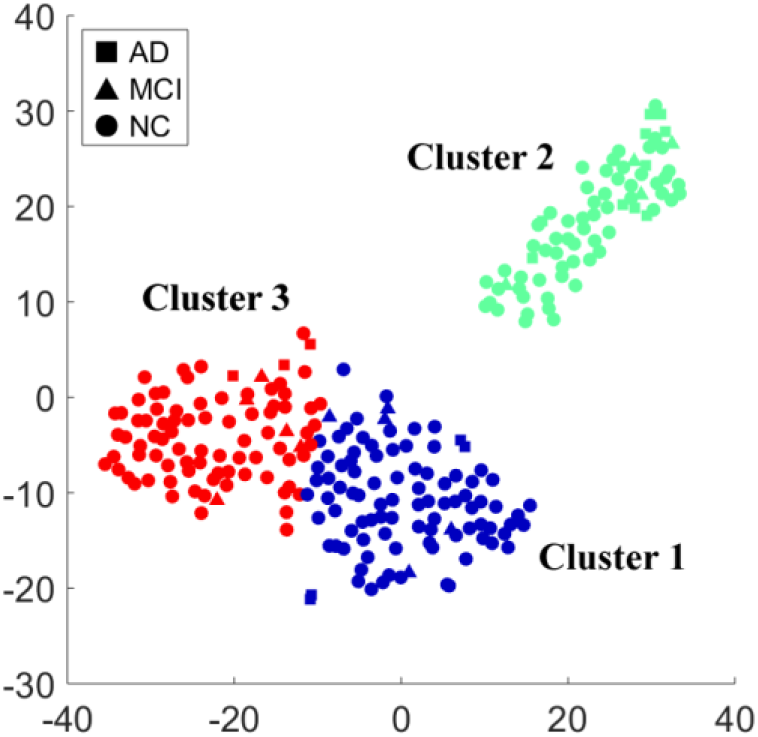
The distribution of samples labeled “AD”, “MCI”, and “NC” in three clusters.

The Cluster 2 has a prominent high Aβ deposition in the lateral and medial prefrontal cortices, the lateral temporoparietal regions, and the posterior cingulate cortex. This is consistent with the previously reported “positive Aβ-PET pattern” in AD patients (Nordberg et al., 2010; Palmqvist et al., 2017).

On the contrary, the Cluster 3 has no elevated Aβ throughout the brain, forming a clear “negative” pattern that is consistent with the previously reported brain Aβ deposition in the healthy brain (Palmqvist et al., 2017).

Interestingly, we found an intermediate Aβ deposition level in Cluster 1 that sits in-between Cluster 2 and Cluster 3, with elevated Aβ deposition in the in the white matter but still low Aβ load in the gray matter (except the frontal area, which has slightly increased gray matter Aβ load compared to the other brain regions).

The three clusters were thus regarded as three Aβ-PET phenotypes in the AD continuum and will be used as labels in the following machine learning-based classification.

### 3.2 Relationship between Aβ-PET phenotypes and other clinical variables

In Fig. 4, we plotted the samples labels (“AD”, “MCI”, or “NC”) in the 2D embedding of the Aβ-PET clustering results. It is clear that our detected three Aβ-PET phenotypes based on the Aβ spatial deposition in the brain are not congruent with the conventional neurophysiological test-based “clinical diagnostic labels” (i.e., AD, MCI, and NC). In fact, each Aβ-PET phenotype cluster included all three types of clinical diagnostic labels. The clinical evaluation based on Aβ-PET phenotype and neurophysiological test faces great discrepancy. For example, all three clusters had a similar amount of NC and MCI (with the Aβ positive Cluster 2 having slightly fewer NC), and all three clusters contained AD subjects, except the Cluster 2 (Aβ positive) had more AD subjects than clusters 1 and 3 (Table 1). Statistical comparison showed that the differences in the clinical diagnostic labels had a marginal significance between Cluster 2 and Cluster 1, as well as between Cluster 2 and Cluster 3 (Table 1).

**Table 1.** Comparison of the five clinical variables among the three Aβ-PET phenotypes.

| Clinical variables | Descriptive statistics |  |  | Main comparison among 3 groups (*p* values) | Pair-wise *post-doc* comparisons (*p* values) |  |  |
| --- | --- | --- | --- | --- | --- | --- | --- |
|  | Cluster 1 | Cluster 2 | Cluster 3 |  | Cluster 1 vs. 2 | Cluster 1 vs. 3 | Cluster 2 vs. 3 |
| Gender | 28 / 69 <sup>a</sup> | 38 / 36 | 49 / 38 | 0.000 <sup>1</sup> | 0.003 | 0.000 | 0.528 |
| Age | 66.3 ± 8.0 <sup>b</sup> | 72.4 ± 5.8 | 64.1 ± 8.9 | 0.000 <sup>2</sup> | 0.000 | 0.218 | 0.000 |
| MMSE | 29.0 ± 2.5 <sup>b</sup> | 28.2 ± 2.4 | 29.5 ± 0.7 | 0.000 <sup>2</sup> | 0.105 | 0.177 | 0.000 |
| APOE | 75 / 20 / 2 <sup>c</sup> | 32 / 33 / 9 | 66 / 19 / 2 | 0.000 <sup>2</sup> | 0.000 | 0.994 | 0.000 |
| Clinical diagnostic labels | 88 / 5 / 4 <sup>d</sup> | 60 / 4 / 10 | 79 / 5 / 3 | 0.120 <sup>1</sup> | 0.085 | 1.000 | 0.068 |

Superscripts indicate the meaning of numbers in various clinical variables and the methods used for comparison. a: The numbers of male and female, b: The means and standard deviations (SDs) of age or MMSE, c: The numbers of 1, 2, and 3 in the statuses of APOE genotype, d: The numbers of NC, MCI, and AD in clinical diagnostic labels, 1: Groups comparison was performed using Chi-square test, 2: Groups comparison was performed using one-way analysis of variance (ANOVA) test.

In addition to comparing with the conventional clinical labels based on neurophysiological tests, we further examined the relationship between the three Aβ-PET phenotypes and other four clinical variables, including gender, age, Mini-Mental State Examination (MMSE), and the status of polymorphism in the apolipoprotein E gene (APOE). We used 1, 2, and 3 to represent the statuses of the polymorphism in the APOE genotype (1 for E2/E2, E2/E3, E3/E3 – low risk; 2 for E2/E4, E3/E4 – medium risk; 3 for E4/E4 – high risk (Farrer et al., 1997)). The results were summarized in Fig. 5 and Table 1.

**Fig. 5.**
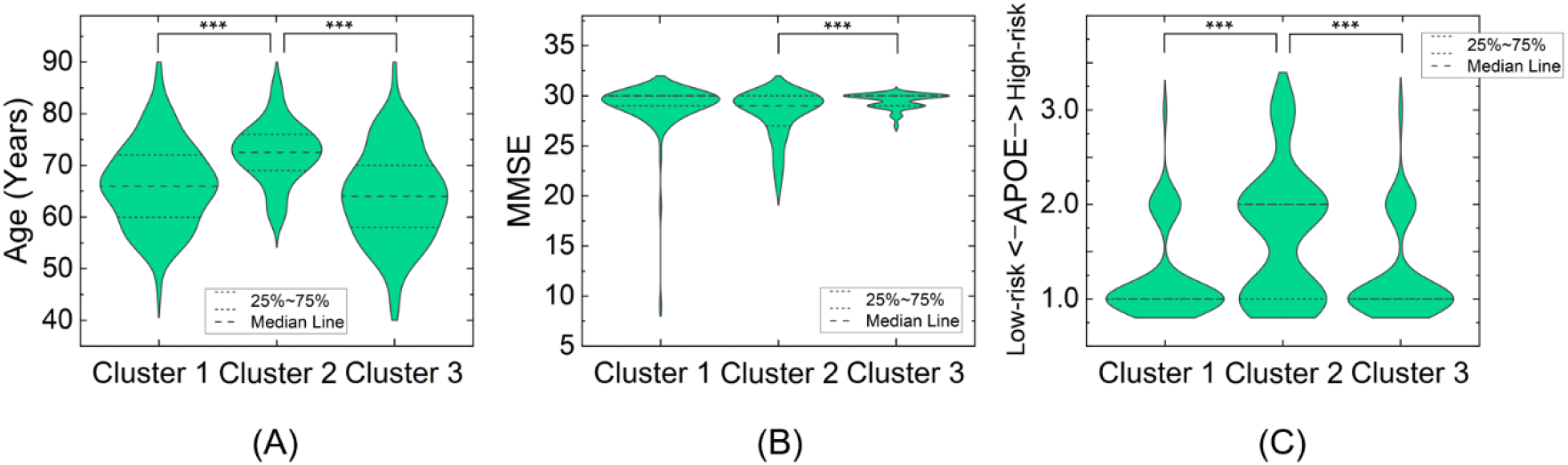
Differences in age (A), MMSE (B), and APOE genotypic status (C) among the three clusters (*** *p* < 0.001, *post hoc* analysis). We used 1, 2, and 3 to represent the statuses of the polymorphism in the APOE genotype (1 for E2/E2, E2/E3, E3/E3 – low risk; 2 for E2/E4, E3/E4 – medium risk; 3 for E4/E4 – high risk).

Gender is a risk factor for AD. We found significant gender differences (*p* < 0.001, Chi-square test) among the three Aβ-PET phenotypes, with the Cluster 1 having a significantly higher female-to-male ratio than those in the Cluster 2 (*p* < 0.01, *post-hoc* analysis) and the Cluster 3 (*p* < 0.001, *post-hoc* analysis).

Age is another key risk factor for AD. We found significant age differences (*p* < 0.001, *F*_2,255_ = 24.099, one-way analysis of variance (ANOVA) test) among the three Aβ-PET phenotypes (Fig. 5A), where the subjects in the Cluster 2 (positive Aβ) are significantly older (mean age: 72.4) than those in the other two clusters (*p* < 0.001 in the *post-hoc* comparisons). The subjects’ age in the clusters 1 (mean age: 66.3) and 3 (mean age: 64.1) did not differ significantly.

MMSE reflects general cognitive ability. We found significant cognitive ability differences (*p* < 0.001, *F*_2,255_ = 8.096, ANOVA test) among the three Aβ-PET phenotypes, with the *post-hoc* comparison indicating that the subjects in the Cluster 2 (positive Aβ) had a lower cognitive ability (*p* < 0.001, *post-hoc* analysis) compared to those in the Cluster 3 (negative Aβ) (Fig. 5B).

APOE is also a major genetic risk factor for AD, with higher risk to AD in carriers of APOE ε4 allele and lower risk in carriers of APOE ε2 allele compared to the carriers of APOE ε3 allele (Yamazaki et al., 2019). We found significant APOE differences (*p* < 0.001, *F*_2,255_ = 16.499, ANOVA test) among the three Aβ-PET phenotypes, with the subjects in the Cluster 2 (positive Aβ) having a significantly higher risk than those in the clusters 1 and 3 (*p* < 0.001, *post-hoc* analysis with ANOVA test, Fig. 5C). It indicated that APOE ε4 allele can significantly increase the expression of Aβ in the brain, consolidating the previous findings (Huang et al., 2017).

In summary, the Aβ-PET phenotype 2 included older subjects with lower cognitive functions and higher genetic risks to AD, while the Aβ-PET phenotype 3 consisted of younger subjects with better cognitive performances and lower genetic risks to AD. Except for having more females (a high-risk gender compared to male), the Aβ-PET phenotype 1 with intermediate Aβ deposition had subjects’ age and MMSE in-between of the clusters 2 and 3, suggesting an intermediate stage between Aβ negative and Aβ positive.

### 3.3 Successful prediction of Aβ-PET phenotypes using brain functional networks

The means and standard deviations (SDs) of the fMRI-to-PET prediction (two-class classification) accuracy, sensitivity, and specificity are listed in Table 2. In all the tasks, fMRI-to-PET prediction with both PC-based and dHOFC-based brain functional networks outperformed those with single brain network. The averaged prediction accuracy was 78.8% across three tasks when both types of brain networks were used, significantly higher than 72.7% when dHOFC was used, and 70.2% when PC was used. Such a result suggests the effectiveness of our proposed fMRI-to-PET prediction method using two types of brain networks.

**Table 2.** Aβ-PET phenotype prediction based on GCN on the multi-level FC networks.

| FC network used | Groups | Accuracy (%) | Sensitivity (%) | Specificity (%) |
| --- | --- | --- | --- | --- |
| PC | Phenotype 1 vs. 2 | 71.4 (3.0) | 74.0 (4.4) | 69.0 (3.6) |
|  | Phenotype 2 vs. 3 | 70.1 (2.0) | 70.6 (6.2) | 70.2 (3.3) |
|  | Phenotype 1 vs. 3 | 69.2 (2.3) | 68.5 (5.5) | 70.0 (5.4) |
|  | Average | 70.2 | 71.0 | 69.7 |
| dHOFC | Phenotype 1 vs. 2 | 72.6 (1.7) | 66.1 (3.1) | 77.0 (1.9) |
|  | Phenotype 2 vs. 3 | 73.0 (2.8) | 72.3 (6.0) | 74.3 (3.8) |
|  | Phenotype 1 vs. 3 | 72.4 (2.1) | 69.6 (6.4) | 73.9 (5.5) |
|  | Average | 72.7 | 69.3 | 75.1 |
| PC+dHOFC | Phenotype 1 vs. 2 | **80.7 (2.7)** | **79.8 (7.3)** | **80.1 (7.7)** |
|  | Phenotype 2 vs. 3 | **79.1 (2.0)** | **77.3 (6.2)** | **80.9 (4.7)** |
|  | Phenotype 1 vs. 3 | **76.7 (1.3)** | **77.2 (3.1)** | **75.3 (3.7)** |
|  | Average | **78.8** | **78.1** | **78.8** |

Values indicate the means (SDs) of accuracy, sensitivity, and specificity from five-fold cross-validation. PC: Classification with Pearson’s correlation-based brain functional network only, dHOFC: Classification with dynamics-based high-order functional connectivity network only, PC + dHOFC: Joint classification based on both PC and dHOFC networks.

## 4. Discussion

In this study, we explored the feasibility of detecting typical Aβ-PET phenotypes through an unsupervised clustering algorithm, with three phenotypes revealed, corresponding to different cognitive statuses, ages, and risk genotypes. According to the spatial patterns (Fig. 3) and their association with various clinical variables (Fig. 5, Table 1), we believe that the clusters 2 and 3 correspond to Aβ positive and negative, while the Cluster 1 denotes an intermediate Aβ deposition stage in between the clusters 2 and 3.

Of note, the data-driven clustering was based solely on the Aβ-PET patterns without human interference, without consideration of the conventional, neurophysiological test-based clinical diagnosis. Based on the overall level and the spatial distribution of the Aβ deposition (SUVR value), we assumed that the phenotype 3 could be treated as a normal status with overall low Aβ load, and the phenotype 2 could be an classic AD-related status with a heavy Aβ load distributed unevenly in the brain (mainly in the brain regions of the default mode network). As for the phenotype 1, being between phenotype 2 and phenotype 3 in both Aβ load and the association with age, MMSE, and APOE, it could be the intermediate stage between negative Aβ and positive Aβ, a middle stage in the AD continuum. This three-phenotype classification is the first observation in the field, comparing to the traditional binary stratification of the Aβ-PET patterns.

Such a three-way Aβ-PET phenotyping could be of potential clinical values. First, under the Aβ catastrophe hypothesis in AD pathogenesis, we provided a totally automatic Aβ-PET phenotyping without setting any threshold, based purely on the Aβ-PET images. Second, we found out an intermediate pattern between a too rough binarization of the Aβ-PET patterns, providing more flexibility and details for clinical evaluation or intervention. Third, the three-way Aβ-PET phenotyping is able to well fit the gradual AD progression process (i.e., the AD continuum) than the binary Aβ-PET phenotyping by defining an intermediate Aβ-PET phenotype.

We further observed age, gender, and genetic differences among the Aβ-PET phenotypes. Age plays a critical role in AD progression, acting as an independent risk factor of AD (Herrup, 2010). In addition, gender, specifically female, is also an important risk factor of AD (Ferretti et al., 2018). In the United States, about two-thirds of people aged 65 and over with AD dementia were women (Nebel et al., 2018). We also observed the association between Aβ-PET deposition and the gene variation (i.e., APOE ε4) (Ishii & Iadecola, 2020). In the phenotype 2, age, MMSE, and genetic risk factor were found to be significantly different compared to the other two Aβ-PET phenotypes. More interestingly, the phenotypes 1 and 3 exhibited similar ages, MMSE scores, APOE types, and even conventional cognitive-test-based clinical labels, indicating they are quite close in terms of the stage of AD progression. The above four factors were not likely to contribute in the stratification between these two phenotypes. The only differences we observed between these two phenotypes is gender, with more females (risky gender) in the phenotype 1 than phenotype 3. We believe that this is not a trivial gender differentiation between the phenotypes 1 and 3, but rather an indication of gender risk in AD, because the gender ratio in the phenotype 3 is quite close to one, with 49 (M) and 38 (F) included. If gender is the sole reason in this classification, the gender ratio in the phenotype 3 could be much larger than one. Therefore, we consider that the phenotype 1 could be a preposition period of Aβ positive.

Compare to the conventional cognitive test-based AD diagnosis strategy, based on Aβ-PET phenotyping, one could better identify potential AD patients based on the Aβ-PET imaging years before the cognitive symptom appears. The inconsistency between Aβ-PET-based phenotyping and the cognitive test-based phenotyping is well reflected in Fig. 4. Such an inconsistency could be caused by many reasons. First, dementia could be caused by many other reasons instead of AD. These non-AD related reasons include other physical diseases, head trauma, virus infection, and so on (Mendez, 2017; Biessels & Whitmer, 2020). Relying only on cognitive test may lead to a false positive AD diagnosis in the clinical practice, which may explain the existence of “AD patients” in the phenotype 3. Second, cognitive test-based diagnosis may lag behind Aβ-PET deposition in the brain (Jack, C.R., Jr. et al., 2010). It has been suggested that the abnormality in the brain Aβ deposition appears more than 10 years before the cognitive abnormality or the “MCI status” (Bateman et al., 2012). This could lead to false negatives and the clinical intervention could be late. Such a reason may lie in the fact that there were many “NC” and “MCI” subjects in the phenotype 2.

In this study, we did not stop at the identification of the three Aβ-PET phenotypes. Importantly, we proposed a novel algorithm to “predict” brain Aβ-PET phenotypes using a non-invasive, easy-to-acquire, low-cost, resting-state fMRI only. Our study demonstrated *not only* the existence of distinctive Aβ-PET phenotypes in the AD continuum *but also* the predictability of such Aβ-PET phenotypes with MRI technique with the help of deep graph learning. We, for the first time, demonstrate the effectiveness of using various brain functional networks derived from fMRI to approximate Aβ-PET-based AD diagnosis. The reason of such a success roots deeply in the physiological mechanism of the BOLD fMRI and the fMRI-derived brain functional connectivity.

BOLD fMRI indirectly detects neural activity and partially reflects and relies on the normal association between the neuronal activity and the blood vessel response. AD-related Aβ deposition may not only cause neuron death, reducing the neuronal activity, but also impair the healthy neurovascular coupling (Hsu et al., 2010). Thus, AD-related Aβ deposition may cause BOLD fMRI changes, which will further lead to functional connectivity changes. Second, AD-related Aβ deposition could make signal transmission less efficient between neurons through axis, due to neural degeneration and necrosis (Reiss et al., 2018) as well as neuronal inflammation (Welikovitch et al., 2020). This could be well captured by fMRI-based functional connectivity. Third, AD may cause reduced brain metabolism and cerebral blood flow, which may be further coupled with fMRI signal and lead to changes in functional connectivity.

Please be noted that our study is different from the previous fMRI-based AD diagnosis whose essence was to predict *cognitive ability*, rather than *Aβ deposition* as a necessary gold standard of AD, using fMRI. Although the previous studies have indicated the success of cognitive ability-based prediction, it is not an essential AD pathology prediction. For example, deep learning technique was used in the previous study with 3D CNN framework learning from fMRI metric maps (i.e., 3D FC maps) to detect MCI and achieved 74.23% accuracy (Kam, T.-E. et al., 2018). However, there are still problems hindering more accurate individualized diagnosis. First, the cognitive test-defined MCI could be a quite heterogeneous group with a small part of them caused by AD, and many others caused by other reasons. As for the further treatment plan for the detected MCI subjects, since such a detection strategy could not identify the reason of MCI, it is difficult for the doctors to conduct suitable intervention to either delay the progression of AD or treat other diseases. Second, predicting cognitive ability using fMRI may be confounded by many influencing factors. For example, doctors’ experience, and patients’ emotional status, cooperation, education level could all affect the result of cognitive ability test. These factors may introduce huge uncertainty in the prediction. However, the prediction of Aβ is more reliable as imaging-based Aβ evaluation is more objective. It can provide the doctors more useful and specific guidance about the Aβ level according to the current AD diagnosis guideline. Third, we argue that the diagnosis accuracy may not be able to further increase if a “clean” MCI group could not be obtained and the cognitive ability could not be accurately quantified. On the other hand, fMRI-based Aβ phenotyping could be further improved in the future.

Compared to the previous MRI-based (with structural MRI predominantly, such as T1-, T2-weightd imaging) and multimodal brain imaging-based (with both structural MRI and multiple PET modalities, including Aβ-PET, tau-PET, FDG-PET) AD diagnosis studies, our study is fundamentally different in terms of how to integrate multimodal images. Previous multimodality studies mainly used Generative Adversarial Networks and representation learning for the diagnosis of AD (Pan, Y. et al., 2018; Zhou, T. et al., 2019). But these studies mainly used MRI to predict missing PET (i.e., image synthesis or data completion), and further used the MRI and the generated PET to jointly predict AD. This type of analysis is on one hand similar to our work (PET prediction), on the other hand, still can only generate missing PET for a few subjects, with most of others having complete MRI and PET. In other words, not like our MRI-based PET prediction, these previous studies actually put MRI and PET in the same position with the same importance. In fact, according to the AD diagnosis guidance (Jack, C.R. et al., 2018), their importance and role in the diagnosis of AD are different. A better approach should be use multimodal MRI to predict PET to achieve the gold-standard-based accuracy in AD prediction. In this sense, the association between structural MRI and PET should be investigated. In our opinion, this association may not as close as that between fMRI and PET, as we have analyzed above.

There is good clinical significance to predict Aβ using fMRI. It only requires significantly less preparing and scanning time (∼ 5-10 minutes) and less cost with less clinical restrictions, and is thus suitable for large-scale AD screening. One may use our method to find out the Aβ positive elderly subjects and treat them as high-risk subjects to conduct a subsequent PET examination. Such an AD screening strategy could be very efficient given the vast availability of 3T MRI scanner in the hospitals and research institutes. Most importantly, compared to clinical evaluation-based screening, our method could identify AD at an even earlier stage, right after abnormal Aβ deposition. Moreover, many post-processing and network construction methods of fMRI can be combined with the currently used two networks in the future to define multi-view brain connectomes at different scales, so as to achieve better prediction accuracy.

In this work, FC networks extracted with different methods reflect different aspects of brain network topologies. The brain network based on PC reflects pairwise interactions between a node and its first neighbors, which is thus node-centric. The dHOFC network, on the other hand, reflects the interactions among time-varying connections (edges) of the brain and provides an edge-centric network (Faskowitz et al., 2020). Our experiment showed that the integration of PC and dHOFC could better capture the effect of the Aβ deposition to the brain networks and predict the Aβ-PET phenotypes in a higher accuracy. Note that the accuracy in differentiating Aβ-PET phenotype 1 from the phenotype 3 achieved 76.7%, which is already comparable to the best reported performance in the previous individualized MCI identification studies (Suk et al., 2017; Li, F. et al., 2018; So et al., 2019), and higher than the reported performance in the early-MCI (the early stage of MCI) identification studies (Kam, T.E. et al., 2020).

There are several limitations in this study. First, we only used Aβ-PET to identify PET phenotypes and use fMRI to predict Aβ PET phenotypes. However, it is reported that tau-PET could also be used to detect AD pathology (Chandra et al., 2019), while FDG-PET has been shown to have supportive role in AD diagnosis (Pan, X. et al., 2021). In the future, we will develop fMRI-based PET phenotype prediction for multiple PET technique for better AD detection. Second, our PET phenotyping will never be the only one AD staging method, and there may be more than one subtyping definitions for different AD stages. For example, there is a symptom-based MCI subtyping method, separating vascular MCI, non-amnestic MCI from amnestic MCI (Salvadori et al., 2016), each subtype of AD having a different evolutionary trajectories (Vogel et al., 2021). Finally, we performed bi-classification among the three phenotypes; a regression-based prediction could be more suitable and helpful for region-wise PET SUVR prediction, which has more details kept for fine-grained AD staging.

## 5. Conclusions

In this work, we demonstrated the effectiveness of a novel early AD detection method that used a non-invasive brain functional imaging technique (fMRI) with advanced deep graph learning on multi-view brain functional networks to differentiate three identified typical and biologically meaningful Aβ-PET phenotypes for better characterzing the AD continuum. Our method indicates a promising future of high-throughput AD screening with less costs and restrictions for subjects at even earlier (preclinical) stage of AD, which could be of great use and clinical value.

## Data Availability

The data used to support the findings of this study are included within the article.

https://www.oasis-brains.org

## CRediT authorship contribution statement

**Chaolin Li:** Data curation, Formal analysis, Visualization, Writing - original draft, Funding acquisition. **Mianxin Liu:** Methodology, Writing - review & editing. **Jing Xia:** Methodology, Writing - review & editing. **Lang Mei:** Software. **Qing Yang:** Writing - review & editing. **Feng Shi:** Writing - review & editing. **Han Zhang:** Conceptualization, Methodology, Supervision, Writing - review & editing. **Dinggang Shen:** Conceptualization, Methodology, Supervision, Project administration, Writing - review & editing.

## Acknowledgments

This work was supported by the Guangzhou Science and Technology Plan Project (No. 202102010495), the ShanghaiTech Startup funds, and the National Key Scientific Instrument Development Program (No. 82027808). Han Zhang was supported by Shanghai Zhangjiang National Innovation Demonstration Zone Special Funds for Major Projects (“Human Brain Research Imaging Equipment Development and Demonstration Application Platform” (ZJ2018-ZD-012)).

